# AI-Derived ECG Age Gap as a Digital Biomarker for Cardiovascular Risk: External Validation in Hospital and Community-Based Prospective Cohorts

**DOI:** 10.64898/2026.03.24.26349186

**Authors:** Shun Huang, GuangKun Nie, Hongmin Liu, Donglin Xie, Jun Li, Gongzheng Tang, Chenyang He, Qinghao Zhao, Deyun Zhang, Jingyuan Jiang, Hao Zhang, Zhengkai Xue, Guodong Wang, Shuohua Chen, Liming Lin, Junqing Xie, Qi Xu, Shouling Wu, Kangyin Chen, Luxia Zhang, Shenda Hong

## Abstract

Cardiovascular diseases remain the leading cause of global mortality, and early risk stratification is critical for improving prognosis. Artificial intelligence–derived electrocardiography (AI-ECG) provides a promising approach to derive cardiac biological age as a non-invasive digital biomarker. This study developed an AI-ECG framework based on the ECGFounder foundation model to quantify cardiac biological aging and predict cardiovascular risk. A total of 67,824 ECGs from 63,512 UK Biobank participants were included. The model was trained on a development cohort of healthy individuals (n = 26,871), and evaluated in an independent clinical evaluation cohort (n = 40,953). Cox proportional hazards models were used to assess the association between the AI-ECG age gap and MACCE, as well as other secondary outcomes. External validation was further conducted in an inpatient cohort from Tianjin Medical University Second Hospital (n = 55,860) and the Kailuan community-based prospective cohort (n = 27,065). The model demonstrated good agreement between predicted and calendar age in the development cohort (r = 0.55; MAE = 5.12 years). In the clinical cohort, after adjusting for clinical comorbidities, each 1-year increase in the age gap was associated with a significant 13% higher risk of MACCE (HR = 1.13, 95% CI: 1.11–1.14). Individuals with an overestimated age gap (> 6 years) exhibited substantially elevated risks of MACCE (HR 4.51) and other major cardiovascular outcomes, whereas those with an underestimated age gap ( <− 6 years) showed a significantly lower risk of MACCE (HR 0.46) alongside protective effects across other outcomes. In the external cohort, the AI-ECG age gap remained significantly associated with multiple downstream cardiovascular and cardiometabolic outcomes. The AI-ECG age gap effectively quantifies occult accelerated cardiac aging. As a non-invasive digital biomarker, it has substantial potential for cardiovascular risk stratification in broad populations.

## INTRODUCTION

Cardiovascular diseases (CVDs) remain the leading cause of mortality and reduced quality of life worldwide^1,2^. According to World Health Organization (WHO) data, approximately 19.8 million people globally died from CVDs in 2022, accounting for about 32% of all global deaths^3^, posing a continuous and severe challenge to public health systems^4^. Although early risk identification and timely intervention are crucial for improving patient prognosis, traditional clinical screening methods are often limited by access to medical resources or rely on overt pathological features^5–7^. As an essential diagnostic tool for CVDs, the electrocardiogram (ECG) has become an ideal initial screening instrument due to its advantages of low cost, non-invasiveness, and high accessibility^8^. However, traditional ECG interpretation primarily relies on the visual assessment of overt waveform characteristics by cardiologists. While this rule-based approach offers clear clinical interpretability, it frequently fails to detect subtle fluctuations and complex patterns embedded within the signals, making it difficult to capture early, occult functional decline in the cardiovascular system^9,10^.

In recent years, the rapid development of artificial intelligence (AI) technologies has provided novel perspectives to extract deeper insights from medical data^11–13^. In particular, deep learning algorithms have been proven capable of identifying high-dimensional, nonlinear features in ECG signals that are imperceptible to human vision^8,14–16^. Studies indicate that the heart is not merely a mechanical pump for blood circulation, but also a sensitive sensor of the body’s overall physiological state, as its electrophysiological activities encode comprehensive information regarding human metabolism and autonomic nerve regulation^17–19^. Therefore, utilizing AI for the deep decoding of ECG signals can not only diagnose specific cardiac pathologies but also quantify an individual’s biological aging, thereby driving a paradigm shift in medical applications from disease diagnosis to health assessment.

Based on this concept, and inspired by digital biomarkers and electrocardiomics^20–22^, the concept of AI-ECG age has been proposed, which refers to the cardiac biological age inferred directly from raw ECG waveforms to characterize the overall functional reserve of the cardiovascular system^23–25^. In an ideal state of health, individuals’ AI-ECG age should roughly align with their calendar age. However, for individuals with CVDs or those in a sub-optimal health state, accumulated physiological damage leads to the remodeling of cardiac electrophysiological characteristics^26^, manifesting as an AI-ECG age significantly higher than the calendar age. This age gap represents more than a simple numerical deviation, but quantifies the degree of accelerated cardiac aging relative to calendar age and serves as a predictive biomarker reflecting an individual’s cardiovascular health trajectory.

To evaluate the clinical value and generalizability of AI-ECG age gap as a digital biomarker, this study adopted a multi-cohort design integrating model development, internal outcome assessment, and external validation. We developed an AI-ECG age prediction model in the UK Biobank^27^ using ECGFounder^14^ as the pre-trained backbone with full fine-tuning, and assessed whether the derived AI-ECG age gap was associated with MACCE and specific cardiovascular outcomes, including coronary heart disease, heart failure, stroke, and atrial fibrillation. To further examine its relevance across real-world settings, we validated the calibrated ECG-age gap in two independent Chinese cohorts. The inpatient cohort from Tianjin Medical University Second Hospital was used to assess existing disease burden, whereas the Kailuan community-based prospective cohort was used to evaluate future incident clinical events. In addition, we extended the outcome scope to cardiometabolic conditions, including hypertension and diabetes, to explore whether AI-ECG age gap could provide complementary information on broader cardiovascular-metabolic health. Through this design, we aimed to validate AI-ECG age gap as an outcome-anchored ECG-derived digital biomarker for cardiovascular aging and risk stratification.

## Methods

### Data Sources and Processing

#### Data Sources

This study utilized three independent databases. The primary dataset was derived from the UKB, a large-scale prospective biomedical database that recruited over 500,000 participants from across the United Kingdom, providing rich genetic, physiological, and medical record data. In addition, an inpatient cohort from Tianjin Medical University Second Hospital between 2022 and 2025 and the Kailuan community-based prospective cohort between 2018 and 2022 were included for external validation. The UKB cohort includes multiple assessment phases, comprising an initial assessment (2006-2010) and subsequent repeat assessments and imaging visits, enabling long-term tracking of participants’ health trajectories. In this study, raw waveform data of standard 12-lead ECGs were extracted by parsing the XML format files provided by the UKB, encompassing a total of 67,824 ECG records from 63,512 participants. The corresponding calendar age was calculated from the birth year (Field 34) and birth month (Field 52) provided by the UKB, with the birth date uniformly set to the 15th of the respective month.

#### ECG Signal Preprocessing

To eliminate environmental noise during signal acquisition and extract high-quality electrophysiological features, all ECG signals sequentially underwent a standardized three-step filtering process. First, a 50 Hz notch filter was applied to eliminate powerline electromagnetic interference. Second, a 0.67–40 Hz bandpass filter was used to retain relevant ECG waveforms and filter out high-frequency noise. Finally, a median filter was utilized to remove baseline wander. Subsequently, all signals were subjected to Z-score normalization to eliminate inter-individual baseline differences and accelerate the convergence of subsequent deep learning models.

#### Study Population and Data Splitting

The data flow and screening process of the study population are illustrated in Figure 1, including both internal and external cohorts. To construct a precise AI-ECG age prediction model and validate its clinical prognostic value, we divided the UKB dataset into a Development Cohort and a Clinical Evaluation Cohort. For the Development Cohort, we applied strict exclusion criteria at the participant level. (1) Participants with multiple ECG records due to multiple assessment visits were excluded to ensure the integrity of labels of the training data and prevent data leakage. (2) Participants already diagnosed with circulatory system diseases (defined based on UKB Category 2409), diabetes, or dyslipidemia at the time of ECG recording were excluded. This ensured that the model was trained on physiological signals from healthy individuals, thereby better capturing the electrophysiological characteristics of the natural aging process. After the aforementioned screening, the remaining healthy participants constituted the final Development Cohort and were randomly split in a 7:1:2 ratio into a Training Set, a Validation Set for hyperparameter tuning, and a Test Set for internal performance evaluation. In the Clinical Evaluation Cohort, to validate the predictive efficacy of the AI-ECG age gap as a digital biomarker in a broader population, participants excluded from the development cohort under criterion (2) due to pre-existing cardiovascular or metabolic diseases were directly included in this cohort.

**Figure 1:**
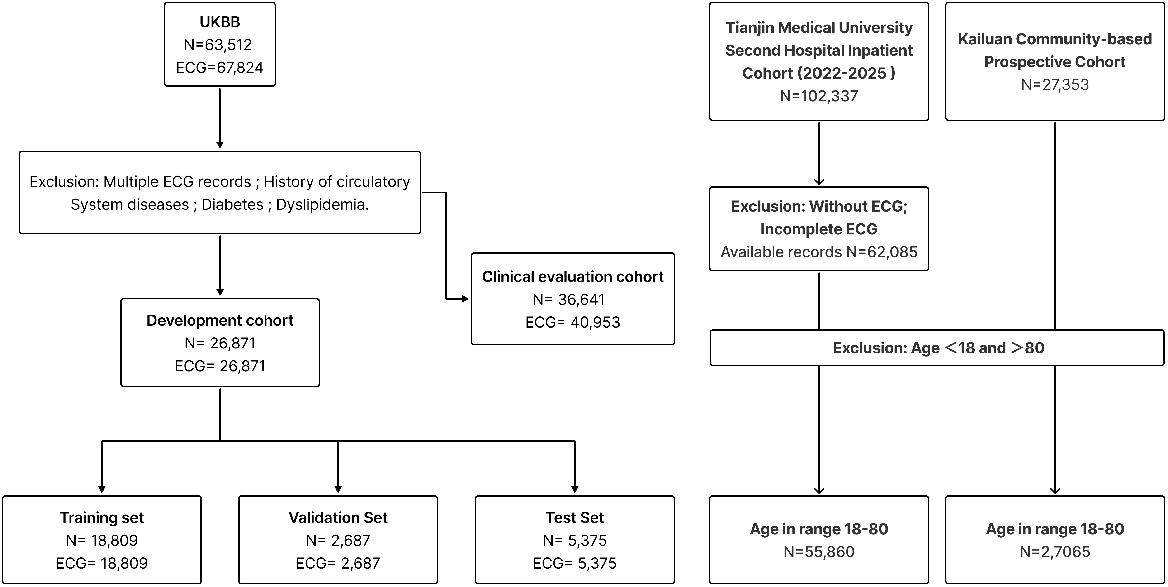
Flowchart of study population screening, model development, internal clinical evaluation, and external validation. Participants from the UK Biobank were screened and divided into a development cohort for AI-ECG age model training and an internal clinical evaluation cohort for prognostic assessment. In addition, this study included an inpatient cohort from Tianjin Medical University Second Hospital and the Kailuan community-based prospective cohort to evaluate model generalizability and downstream clinical associations.

#### Ethical Statement

The UKB obtained ethical approval from the North West Multi-centre Research Ethics Committee (REC reference: 21/NW/0157). All participants included in the UKB provided informed consent for their data to be used in health-related research. This study was conducted under UKB application number 90018. The external validation studies were approved by the Biomedical Ethics Committee of Peking University (Approval No.: IRB00001052-23071), the Ethics Committee of the Second Hospital of Tianjin Medical University (Approval No.: KY2025K386), and the Ethics Committee of Kailuan General Hospital (Approval No.: [2006] No. 5). The requirement for informed consent was waived because the study involved retrospective analyses of de-identified data.

### Model Development

#### Model Architecture

The AI-ECG age prediction model in this study is based on ECGFounder^14^, a universal ECG foundation model pre-trained on a massive dataset of over ten million ECGs. This large-scale pre-training enables the model to learn universal latent space representations of cardiac electrophysiological activities. The architecture of ECGFounder is built upon RegNet (RegNet-1d)^28,29^, employing a multi-stage design that comprises a series of convolutional blocks with Group Convolutions and Channel-wise Attention Mechanisms. This design allows the model to not only capture the fine temporal fluctuations of ECG signals (temporal dependency) but also effectively integrate the spatial correlations across different leads (spatial dependency). This characteristic is crucial for identifying the subtle electrophysiological remodeling associated with cardiac aging. In this study, we employed a Transfer Learning strategy to conduct Full Fine-tuning on ECGFounder for the age prediction task (Figure 2). Specifically, we initialized the model with pre-trained weights, removed the original linear layer for diagnostic classification, and replaced it with a randomly initialized linear output layer dedicated to the regression task. During the full fine-tuning process, we trained the entire network end-to-end, updating not only the newly added regression layer but also allowing the pre-trained backbone network parameters to undergo adaptive updates, thereby precisely transferring the universal ECG feature space to the cardiovascular physiological aging assessment task. The model inputs were standardized 12-lead ECG signals (sampling rate 500 Hz, duration 10 seconds), and the model weights were updated by minimizing the error between the predicted age and the calendar age.

**Figure 2:**
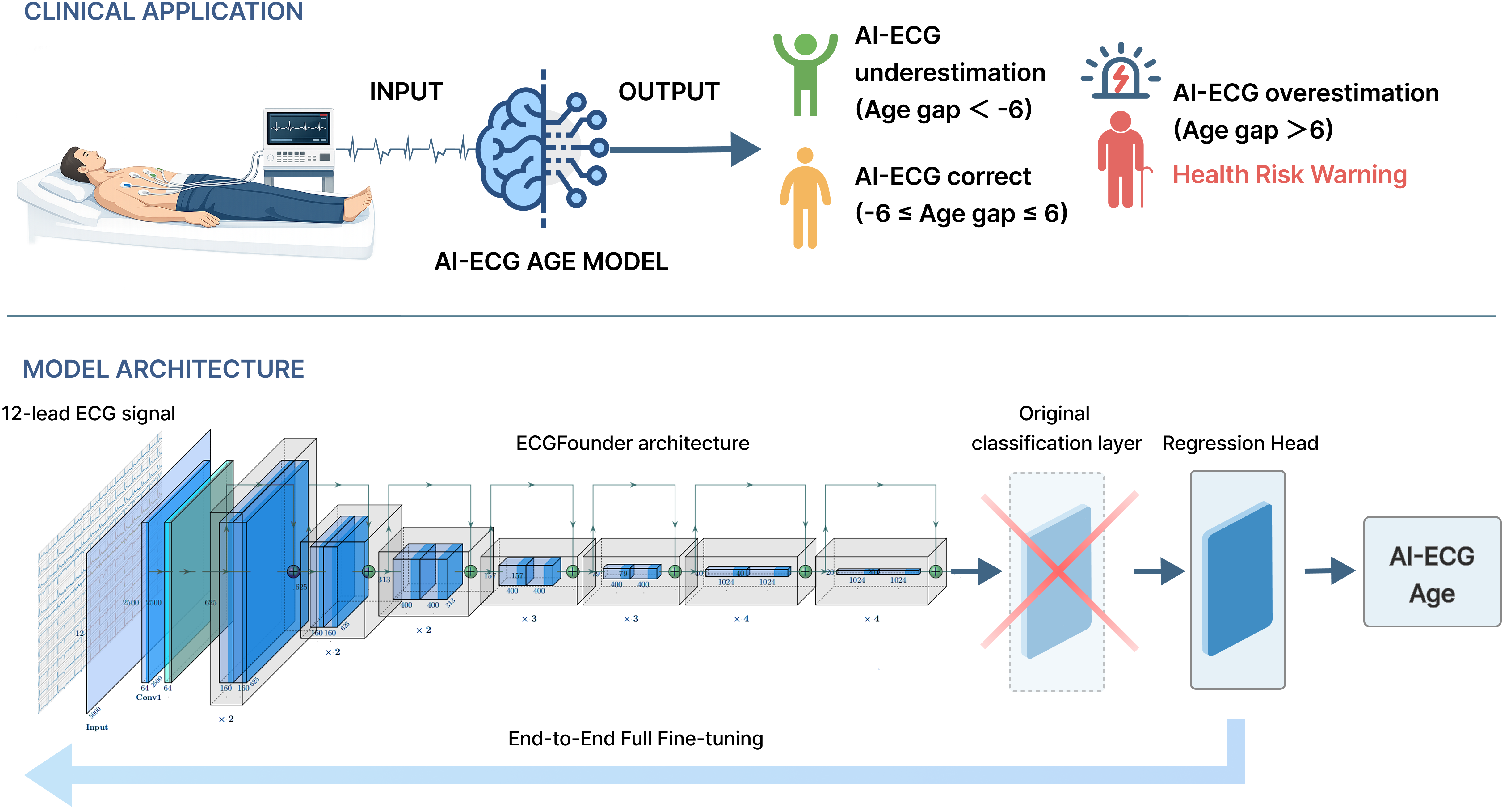
Overall application flowchart and architecture of the AI-ECG age prediction model. The figure presents the clinical application pipeline and the model architecture. The prediction model was built upon the pre-trained ECGFounder and fully fine-tuned to predict calendar age.

#### Experimental Settings

The deep learning models in this study were constructed based on the PyTorch framework and trained on a single NVIDIA GeForce RTX 3090 GPU (24GB VRAM). We utilized the Adam optimizer (with parameters set to *β*_1_ = 0.9, *β*_2_ = 0.999) to perform full fine-tuning of the model for 10 epochs. To ensure training stability, the batch size was set to 256. Regarding learning rate scheduling, the initial learning rate was set to 3 *×* 10^*−*3^, combined with a Cosine Annealing Decay strategy, capping the minimum learning rate during training at 1 *×*10^*−*5^. For model optimization and evaluation, this study directly adopted the Mean Absolute Error (MAE) as both the loss function and the primary evaluation metric, and the model with the lowest validation MAE was selected as the final model during the training process.

#### Model Interpretability

To investigate the internal decision-making mechanisms of the deep learning model when inferring biological age and to enhance its clinical transparency, this study employed saliency maps to visualize a representative case. Specifically, we selected a high-risk subject whose biological age was overestimated by the model. A corresponding saliency map was generated by computing the gradients of the model’s predictive output with respect to the input ECG signals. Regions with higher gradient magnitudes indicate signal segments that contribute most significantly to the model’s final prediction. To improve visualization quality and mitigate the influence of baseline noise, Gaussian smoothing was applied to the original saliency map.

### Outcomes Definition and Clinical Value Assessment

#### Outcomes Definition

To comprehensively evaluate the clinical efficacy of AI-ECG age as a cardiovascular health biomarker, this study prespecified multi-level clinical endpoints within the UK Biobank cohort. (1) Primary Endpoint: Defined as Major Adverse Cardiovascular and Cerebrovascular Events (MACCE), which is a composite endpoint. (2) Secondary Endpoints: Specific major cardiovascular and cerebrovascular events, including Coronary Heart Disease (CHD), Heart Failure, Stroke, and Atrial Fibrillation, were analyzed separately as secondary endpoints. Additionally, Hypertension and Diabetes were included as supplementary cardiometabolic outcomes. The determination of all disease outcomes was based on the International Classification of Diseases, Tenth Revision (ICD-10) codes. For heart failure, CHD, Hypertension, and Diabetes, only the first diagnosed event was recorded, while outcomes for atrial fibrillation and stroke were derived from algorithmically defined criteria.

#### Assessment Strategy

We quantified the degree of individual physiological aging by calculating the AI-ECG age gap, defined as the difference between the AI-predicted age and the actual calendar age (Gap = Age_AI_ *−* Age_Calendar_). To validate its prognostic value, we incorporated the age gap into the analysis using the following two strategies. (1) Continuous Variable Analysis: We treated the AI-ECG age gap as a continuous variable to evaluate the linear or non-linear effects of each 1-unit increase in this metric on the risk of future adverse clinical events. (2) Categorical Variable Analysis: For clinical risk stratification, participants were divided into different risk groups. Using 6 years as the threshold, the population was categorized into an underestimation group (Gap *<−* 6 years), a correct prediction group ( *−*6 years ≤Gap≤ 6 years), and an overestimation group (Gap *>* 6 years). With the correct prediction group serving as the reference baseline, the relative risks of the underestimation and overestimation groups were calculated in comparison to individuals with normal physiological aging.

### External Validation

To assess model generalizability, we further evaluated the AI-ECG age model in two independent external cohorts, including an inpatient cohort from Tianjin Medical University Second Hospital between 2022 and 2025 and the Kailuan community-based prospective cohort between 2018 and 2022. In the retrospective clinical cohort from Tianjin Medical University Second Hospital, 102,337 inpatients were initially screened. After excluding individuals without available ECGs or with incomplete ECG recordings, further screening was performed according to age range and data completeness. The final external validation cohort included 55,860 subjects aged 18–80 years. In the Kailuan community-based prospective cohort, 27,353 participants with available ECG data were matched, and 27,065 subjects aged 18–80 years with complete follow-up information were finally included. The detailed screening process is shown in Figure 1.

Considering that AI-predicted ECG age in external validation cohorts may be affected by regression to the mean and age-related prediction bias, the ECG-age gap was calibrated before downstream clinical association and survival analyses^30,31^. Specifically, AI-predicted ECG age was regressed on chronological age within each external cohort, and the residual from this regression was defined as the calibrated ECG-age gap:

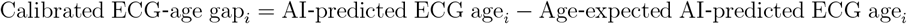

This calibrated ECG-age gap represents the extent to which an individual’s AI-predicted ECG age deviates from the expected value among individuals of the same chronological age. This procedure reduces age-dependent bias caused by compression of the predicted age range or regression to the mean. The calibration used only AI-predicted ECG age and chronological age and did not include any outcome variables. The calibrated ECG-age gap was then standardized as a z score within each external cohort and used for downstream analyses.

For the inpatient cohort from Tianjin Medical University Second Hospital, which was mainly used to assess the cross-sectional association between the ECG-age gap and existing clinical outcomes, logistic regression models adjusted for age and sex were used. Odds ratios (ORs) and 95% confidence intervals (CIs) were reported. For the Kailuan community-based prospective cohort, the primary analysis used Cox proportional hazards models adjusted for age and sex to evaluate the association between the calibrated ECG-age gap and incident clinical events. Hazard ratios (HRs) and 95% CIs were reported. Follow-up time was defined as the period from the baseline ECG examination date to the first occurrence of the target outcome or the end of follow-up. For analyses of specific outcomes, individuals with the corresponding disease at baseline were excluded.

In both external validation cohorts, the primary analysis modeled each 1-standard-deviation increase in the calibrated ECG-age gap as a continuous variable. For categorical analysis, a calibrated ECG-age gap z score greater than 1 was defined as the high calibrated ECG-age gap group, while participants with a calibrated ECG-age gap z score of 1 or lower served as the reference group.

## Statistical Analysis

All data processing and statistical analyses were performed in the R (version 4.2.2) and Python (version 3.11) environments. The main R packages used included survival and survminer for survival analysis, and rms for spline regression modeling. All statistical tests were two-sided, and a P-value < 0.05 was considered statistically significant.

### Descriptive Statistics and Model Performance

For the baseline characteristics of the study population, continuous variables were expressed as mean*±* standard deviation (Mean *±* SD), while categorical variables were reported as frequencies and percentages (*N*, %). To quantify the accuracy and consistency of the AI-ECG age prediction model, we calculated the Pearson Correlation Coefficient (*r*) and Mean Absolute Error (MAE) between the predicted age and the actual calendar age.

### Survival Analysis and Risk Assessment

This study utilized Cox Proportional Hazards Models to evaluate the strength of the association between the AI-ECG age gap and MACCE, as well as various secondary endpoints, with results presented as HRs and their 95% CIs. To intuitively demonstrate the potential non-linear dose-response relationship between the age gap and clinical risks, we applied Restricted Cubic Splines (RCS) for fitting. Based on the risk stratification (underestimation, normal, overestimation), we plotted Kaplan-Meier survival curves to visualize the cumulative event incidence across different groups and employed the Log-rank Test to compare the statistical significance of differences among the groups. Survival time was defined as the period from the participant’s baseline assessment to the occurrence of the endpoint event or the end of the follow-up period (December 31, 2022). For each outcome-specific Cox analysis, participants with the corresponding disease before or at baseline were excluded.

### Covariate Adjustment Strategy

To strictly control the impact of potential confounding factors on the prognostic analysis, we constructed two multivariable adjustment models. Model 1 served as the basic adjustment model and primarily adjusted for demographic characteristics including age, sex, ethnicity, and body mass index. Model 2 further incorporated smoking status and pre-existing clinical comorbidities, including hypertension, diabetes, dyslipidemia, and chronic kidney disease, in addition to the variables included in Model 1.

## Results

### Study Population

In this study, we developed and evaluated the AI-ECG age prediction model based on the UKB dataset. The detailed baseline clinical characteristics of participants in each study cohort are summarized in Table 1. Initially, a total of 67,824 standard 12-lead ECG records from 63,512 participants were extracted from the overall dataset. Based on predefined criteria for independence and health status, the study population was divided into two subsets, the Development Cohort and the Clinical Evaluation Cohort.

**Table 1.** Characteristics of the Study Cohorts.

| Characteristics | Development cohort (N = 26,871) | Evaluation cohort (N = 40,953) |
| --- | --- | --- |
| Age (years) | 63.94 (7.64) | 67.11 (7.69) |
| Sex | N=26,871 | N=40,953 |
| Male | 11,182 (41.61%) | 21,655 (52.88%) |
| Female | 15,689 (58.39%) | 19,298 (47.12%) |
| Race | N=26,865 | N=40,939 |
| White | 25,949 (96.59%) | 39,377 (96.18%) |
| Asian | 346 (1.29%) | 676 (1.65%) |
| Black | 179 (0.67%) | 337 (0.82%) |
| Mixed | 168 (0.62%) | 195 (0.47%) |
| Others | 223 (0.83%) | 354 (0.86%) |
| BMI (kg/m<sup>2</sup>) | 25.69 (4.09) | 27.13 (4.60) |
| Hypertension | 0 (0%) | 20,866 (50.95%) |
| Diabetes | 0 (0%) | 3,948 (9.64%) |
| Dyslipidemia | 0 (0%) | 16,303 (39.81%) |
| Chronic kidney disease | 189 (0.70%) | 1,306 (3.19%) |
| Smoking status | N=20,250 | N=31,864 |
| Non-smoker | 19,544 (96.51%) | 30,807 (96.68%) |
| Current smoking | 706 (3.49%) | 1,057 (3.32%) |
| Heart Failure | 0 (0%) | 530 (1.29%) |
| Atrial fibrillation | 0 (0%) | 2,403 (5.87%) |
| Myocardial infarction | 0 (0%) | 1,652 (4.03%) |
| Stroke | 1 (0.00%) | 793 (1.94%) |
| Coronary heart disease | 0 (0%) | 4,355 (10.63%) |
| MACCE | 1 (0.00%) | 5,017 (12.25%) |
| Follow-up time (years) | 2.86 (2.70) | 2.77 (2.57) |

After excluding individuals with multiple ECG measurement records, as well as those with a history of circulatory system diseases (based on UKB Category 2409), diabetes, or dyslipidemia at the time of measurement, a total of 26,871 healthy subjects (corresponding to 26,871 single ECG records) were included in the Development Cohort. This cohort was subsequently randomly divided into a training set (N = 18,809), a validation set (N = 2,687), and a test set (N = 5,375) in a 7:1:2 ratio. The validation set was used for hyperparameter fine-tuning during model training, while the hold-out test set was used to evaluate the baseline performance of the AI-ECG model in age prediction.

Subjects who were excluded during the screening phase of the development cohort due to a history of cardiovascular or metabolic diseases were all included in the Clinical Evaluation Cohort. This cohort ultimately comprised 40,953 ECG records from 36,641 participants. Encompassing a broad spectrum of samples ranging from sub-optimal health to diagnosed diseases, this cohort was specifically utilized to evaluate the predictive value of the AI-ECG age gap for future cardiovascular clinical outcomes.

### Performance of the AI-ECG Age Prediction Model

To quantify the prediction accuracy of the deep learning model fine-tuned on ECGFounder, we evaluated the consistency between the AI-ECG predicted age and the subjects’ actual calendar age in the model development set (test set), the clinical evaluation cohort, and the external validation cohort. The results demonstrated that in the development cohort consisting of healthy subjects, the AI-ECG predicted age exhibited a linear correlation with the true calendar age, with a Pearson correlation coefficient of 0.55 and a MAE of 5.12 years. In the clinical evaluation cohort, the Pearson correlation coefficient remained comparable at 0.56, while the MAE increased to 5.44 years. The slightly higher MAE in the clinical evaluation cohort may reflect the broader inclusion of subjects with cardiovascular or metabolic diseases, in whom pathological processes may lead to greater deviation between AI-ECG predicted age and calendar age. This supports the subsequent use of the AI-ECG age gap for clinical risk assessment. The scatter plots for the corresponding results are shown in Supplementary Figures S1.

### AI-ECG Age Gap as a Continuous Predictor of Cardiovascular Events

We employed Cox proportional hazards models to evaluate whether the AI-ECG age gap as a continuous variable was independently associated with future adverse cardiovascular events. Detailed results are presented in Table 2. In the basic adjustment model (Model 1, adjusted for age, sex, ethnicity, and BMI), each 1-year increase in the AI-ECG age gap was associated with a significant 12.9% increase in the risk of the primary endpoint, MACCE (HR = 1.129, 95% CI: 1.114–1.145, *p <* 0.001). Furthermore, an increase in this metric was significantly associated with all predefined secondary clinical endpoints included in the analysis (all *p <* 0.001), with specific hazard ratios as follows: CHD (HR = 1.137), Heart Failure (HR = 1.150), Stroke (HR = 1.152), and Atrial Fibrillation (HR = 1.158). Notably, accelerated cardiac aging not only increased the risk of cardiovascular diseases but was also significantly associated with an elevated risk of cardiometabolic diseases such as Hypertension (HR = 1.185) and Diabetes (HR = 1.163). In Model 2, we further adjusted for smoking history and pre-existing comorbidities, including hypertension, diabetes, dyslipidemia, and chronic kidney disease. The results demonstrated that the independent predictive value of the AI-ECG age gap remained highly robust. The hazard ratios for the primary endpoint MACCE and heart failure were 1.126 (95% CI: 1.110–1.142) and 1.149 (95% CI: 1.121–1.177), respectively, and the statistical significance for all follow-up endpoints remained unchanged (all *p <* 0.001).

**Table 2.**
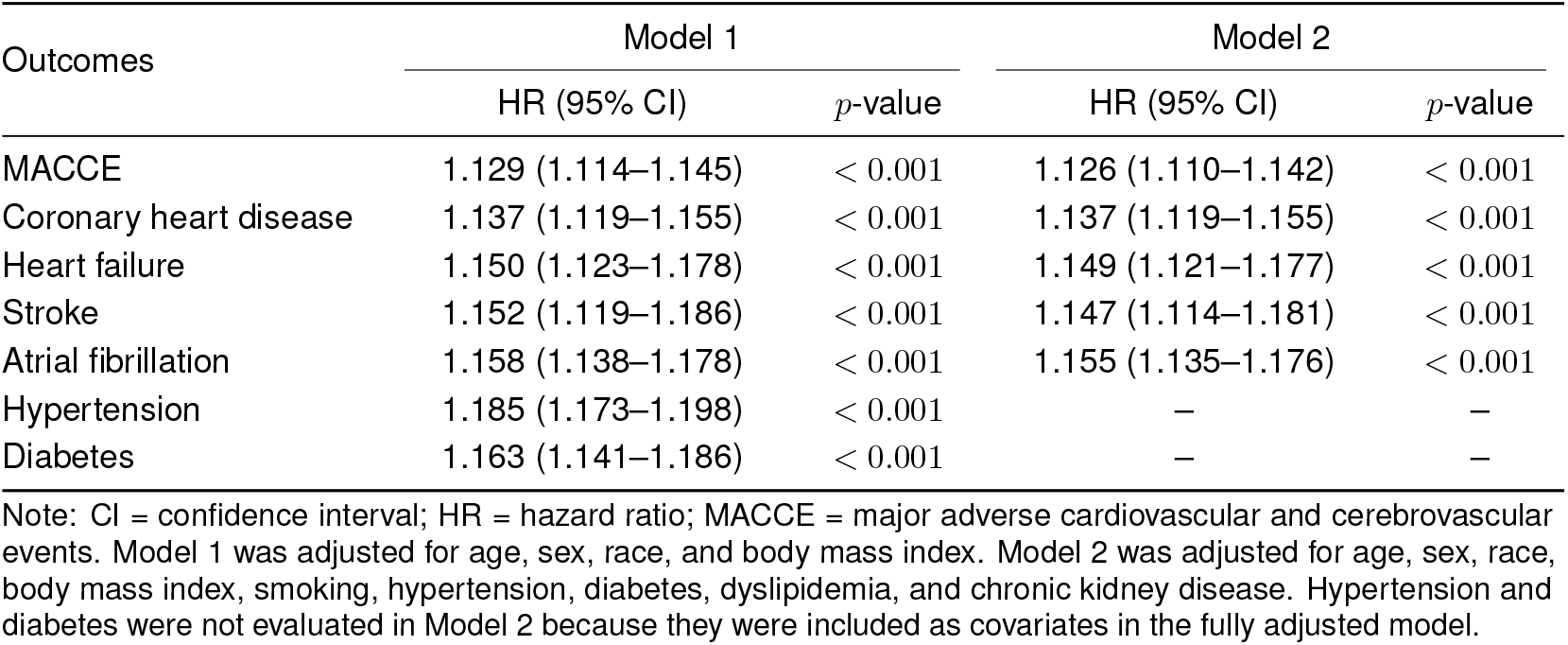
Association of AI-ECG Age Gap as a Continuous Variable with the Risk of Adverse Clinical Outcomes.

To visually illustrate the potential non-linear dose-response relationship between the AI-ECG age gap and the risk of clinical events, we plotted Restricted Cubic Spline (RCS) curves for Model 1, as shown in Figure 3. The RCS curves clearly indicate that as the AI-ECG age gap progressively widened, the relative risk of incident MACCE and various cardiovascular outcomes exhibited a continuous and significant upward trend.

**Figure 3:**
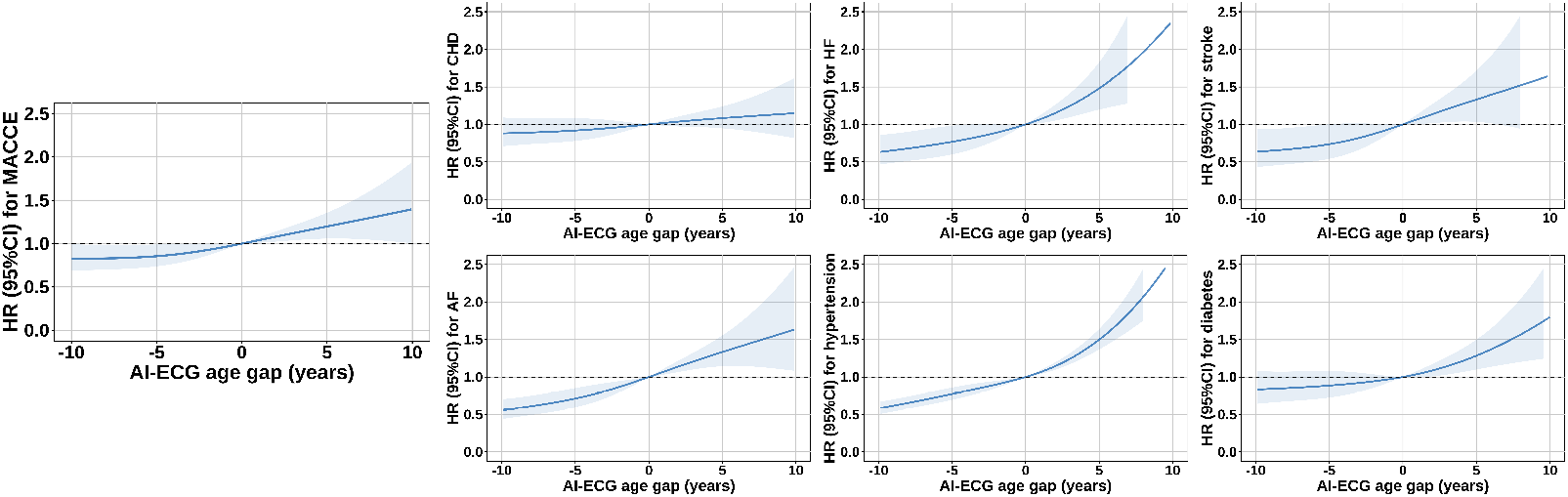
Restricted Cubic Spline (RCS) curves for the association between AI-ECG age gap and the risk of adverse clinical outcomes. The curves (adjusted for Model 1 covariates) demonstrate a continuous non-linear upward trend in the relative risk of MACCE and secondary cardiovascular outcomes as the continuous age gap widens.

### Risk Stratification Based on AI-ECG Age Gap Classification

To evaluate the clinical utility of the AI-ECG age gap for risk stratification, participants were categorized into three groups using a threshold of *±*6 years: Underestimation (Gap *< −*6), Correct Prediction (*−*6 *≤* Gap *≤* 6), and Overestimation (Gap *>* 6). The Correct Prediction group served as the reference baseline in Cox regression analyses to estimate relative risks, with results shown in Table 3.

**Table 3.**
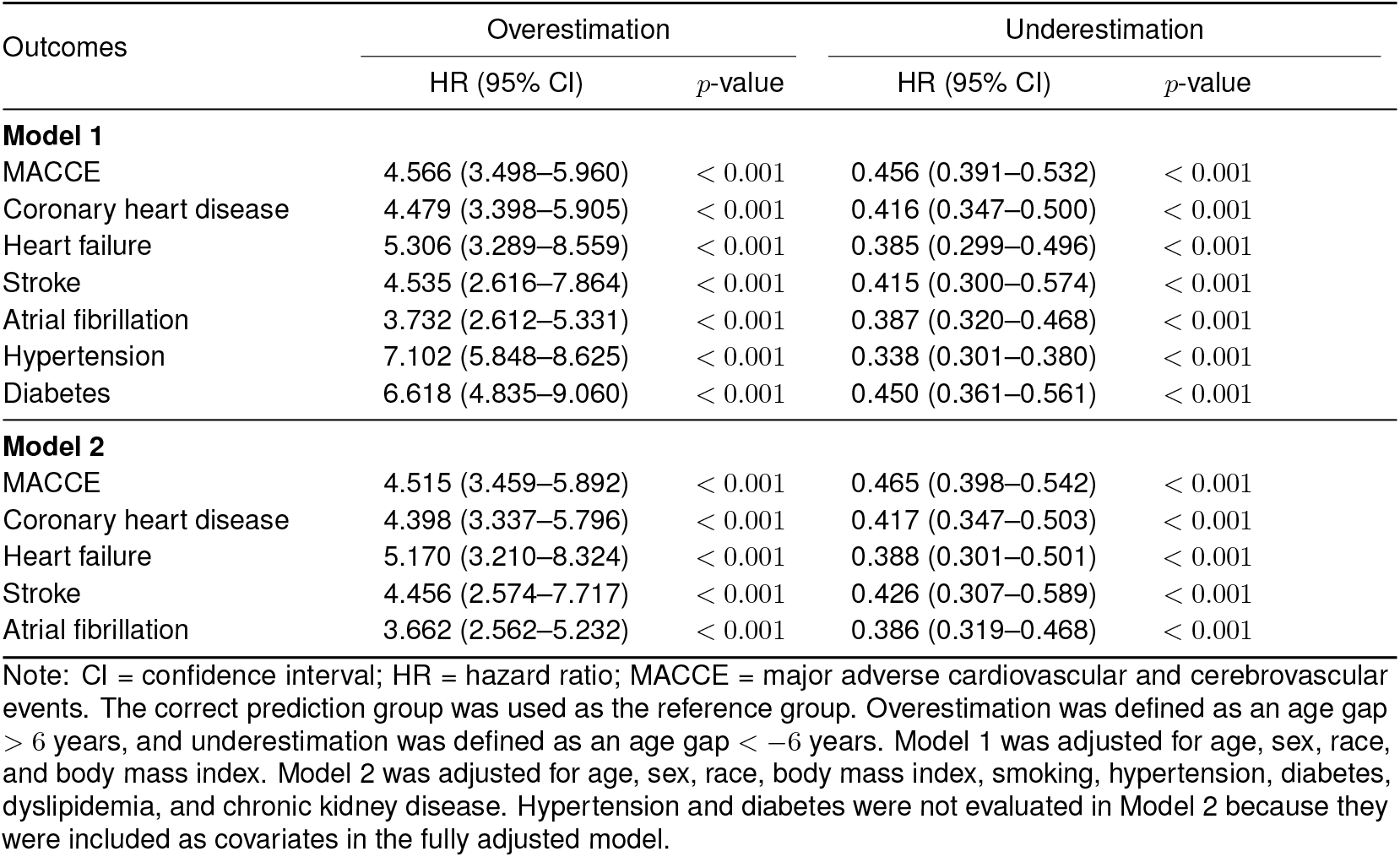
Risk of Adverse Clinical Outcomes Stratified by AI-ECG Age Gap Categories.

| Outcomes | Overestimation |  | Underestimation |  |
| --- | --- | --- | --- | --- |
|  | HR (95% CI) | *p*-value | HR (95% CI) | *p*-value |
| **Model 1** |  |  |  |  |
| MACCE | 4.566 (3.498–5.960) | < 0.001 | 0.456 (0.391–0.532) | < 0.001 |
| Coronary heart disease | 4.479 (3.398–5.905) | < 0.001 | 0.416 (0.347–0.500) | < 0.001 |
| Heart failure | 5.306 (3.289–8.559) | < 0.001 | 0.385 (0.299–0.496) | < 0.001 |
| Stroke | 4.535 (2.616–7.864) | < 0.001 | 0.415 (0.300–0.574) | < 0.001 |
| Atrial fibrillation | 3.732 (2.612–5.331) | < 0.001 | 0.387 (0.320–0.468) | < 0.001 |
| Hypertension | 7.102 (5.848–8.625) | < 0.001 | 0.338 (0.301–0.380) | < 0.001 |
| Diabetes | 6.618 (4.835–9.060) | < 0.001 | 0.450 (0.361–0.561) | < 0.001 |
| **Model 2** |  |  |  |  |
| MACCE | 4.515 (3.459–5.892) | < 0.001 | 0.465 (0.398–0.542) | < 0.001 |
| Coronary heart disease | 4.398 (3.337–5.796) | < 0.001 | 0.417 (0.347–0.503) | < 0.001 |
| Heart failure | 5.170 (3.210–8.324) | < 0.001 | 0.388 (0.301–0.501) | < 0.001 |
| Stroke | 4.456 (2.574–7.717) | < 0.001 | 0.426 (0.307–0.589) | < 0.001 |
| Atrial fibrillation | 3.662 (2.562–5.232) | < 0.001 | 0.386 (0.319–0.468) | < 0.001 |

Note: CI = confidence interval; HR = hazard ratio; MACCE = major adverse cardiovascular and cerebrovascular events. The correct prediction group was used as the reference group. Overestimation was defined as an age gap > 6 years, and underestimation was defined as an age gap < –6 years. Model 1 was adjusted for age, sex, race, and body mass index. Model 2 was adjusted for age, sex, race, body mass index, smoking, hypertension, diabetes, dyslipidemia, and chronic kidney disease. Hypertension and diabetes were not evaluated in Model 2 because they were included as covariates in the fully adjusted model.

In the basic adjustment model (Model 1), the overestimation group exhibited an extremely high risk for adverse prognoses. Specifically, the risk of developing the primary endpoint MACCE in the overestimation group was 4.566 times that of the normal group (HR = 4.566, 95% CI: 3.498–5.960, *p <* 0.001). Regarding secondary outcomes, the risk of organic heart diseases in the overestimation group also showed a precipitous increase, including CHD (HR = 4.479), Heart Failure (HR = 5.306), Stroke (HR = 4.535), and Atrial Fibrillation (HR = 3.732) (all *p <* 0.001). In addition, the risk of incident cardiometabolic diseases in this population surged, with the risks of Hypertension and Diabetes reaching 7.102 times and 6.618 times those of the normal population, respectively. In stark contrast, the underestimation group demonstrated significant protective characteristics across all clinical endpoints. Its risk of MACCE decreased by 54.4% (HR = 0.456, 95% CI: 0.391–0.532, *p <* 0.001), and the risk of developing other cardiovascular outcomes also generally declined by more than 50% (all *p <* 0.001).

In Model 2, the risk stratification results remained highly robust. The relative risk for MACCE in the overestimation group continued to be extremely high (HR = 4.515, 95% CI: 3.459–5.892, *p <* 0.001). Similarly, the risks for other secondary outcomes, including CHD (HR = 4.398), Heart Failure (HR = 5.170), Stroke (HR = 4.456), and Atrial Fibrillation (HR = 3.662), remained significantly elevated. Conversely, the MACCE risk in the underestimation group remained significantly lower than that of the normal population (HR = 0.465, 95% CI: 0.398–0.542, *p <* 0.001), with consistent protective effects observed across all secondary endpoints included in Model 2 (all *p <* 0.001). This demonstrates that the AI-ECG age gap provides an additive risk predictive value independent of traditional clinical comorbidities. Finally, we plotted Kaplan-Meier survival curves (Figure 4) to visually display the cumulative event incidence rates among different AI-ECG age gap groups during the follow-up period. The results revealed that the risk trajectories of the different groups gradually diverged over long-term follow-up; the cumulative incidence curve for the overestimation group exhibited a significantly steeper upward trend, while the curve for the underestimation group remained consistently flatter and situated below that of the normal group. This clear risk stratification pattern further bolstered the clinical utility of the AI-ECG age gap as a potential indicator for risk stratification.

**Figure 4:**
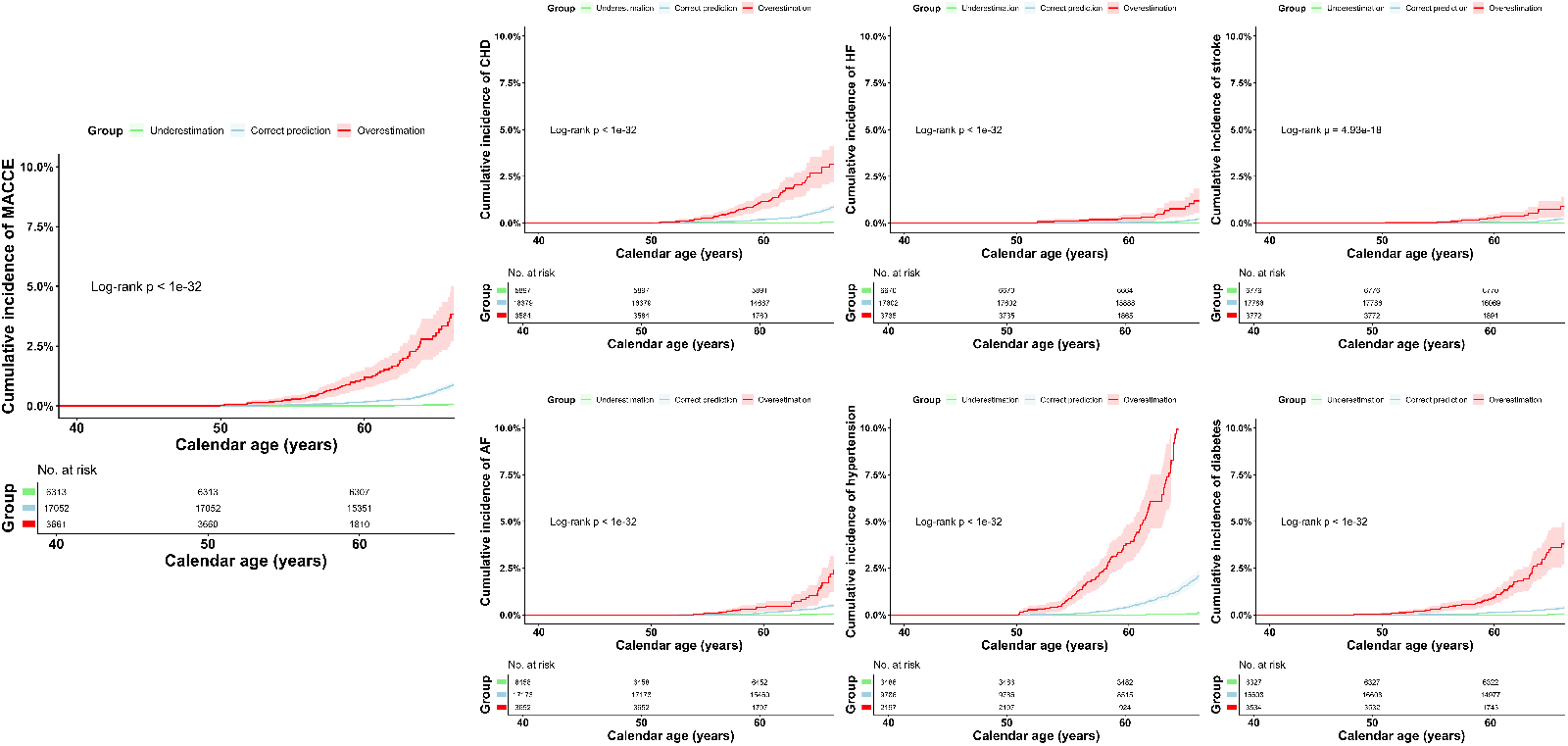
Kaplan-Meier survival curves for cumulative event incidence stratified by AI-ECG age gap. The curves display diverging risk trajectories: the overestimation group (Gap *>* 6 years) exhibits a significantly steeper upward trend for adverse events, whereas the underestimation group (Gap *<* -6 years) demonstrates protective characteristics.

### Clinical Risk Assessment in the External Validation Cohorts

To further validate the clinical relevance of the AI-ECG age gap, we evaluated its associations with adverse clinical outcomes in independent external validation cohorts. As shown in Table 4, in the inpatient cohort from Tianjin Medical University Second Hospital, each 1-standard-deviation increase in the calibrated ECG-age gap was significantly associated with higher odds of MACCE (OR 1.087, 95% CI 1.066–1.108). Similar positive associations were observed for coronary heart disease, stroke, heart failure, atrial fibrillation, and hypertension, with relatively stronger associations observed for atrial fibrillation (OR 1.273, 95% CI 1.231–1.317) and hypertension (OR 1.152, 95% CI 1.132–1.172). In the categorical analysis, individuals with a calibrated ECG-age gap z score greater than 1 also showed a higher burden of clinical disease, particularly for atrial fibrillation (OR 2.208, 95% CI 2.044–2.385), coronary heart disease (OR 1.422, 95% CI 1.356–1.490), and hypertension (OR 1.434, 95% CI 1.368–1.504).

**Table 4.**
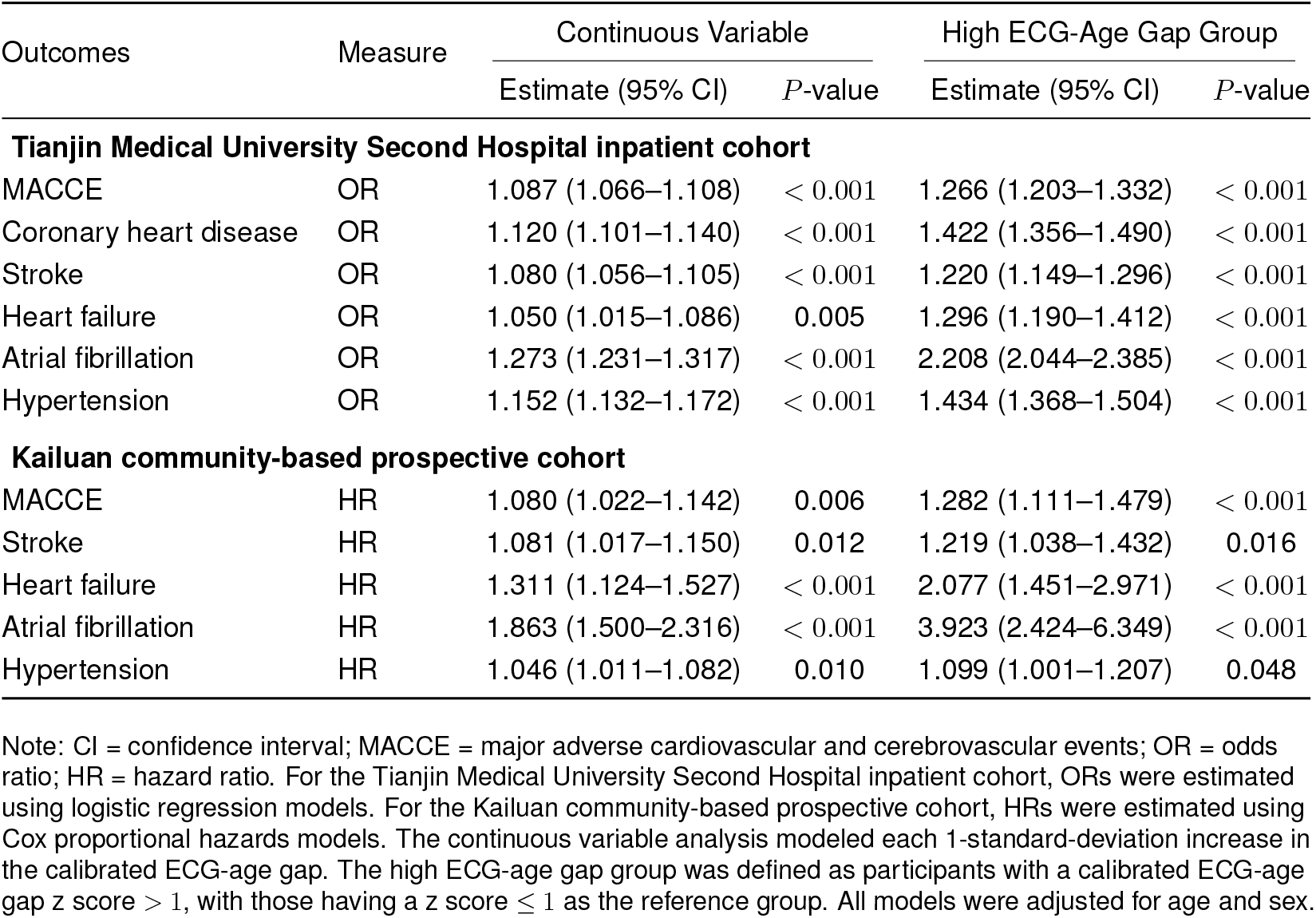
Association of Calibrated ECG-Age Gap with Clinical Outcomes in the External Validation Cohorts.

| Outcomes | Measure | Continuous Variable |  | High ECG-Age Gap Group |  |
| --- | --- | --- | --- | --- | --- |
|  |  | Estimate (95% CI) | *P*-value | Estimate (95% CI) | *P*-value |
| Tianjin Medical University Second Hospital inpatient cohort |  |  |  |  |  |
| MACCE | OR | 1.087 (1.066–1.108) | < 0.001 | 1.266 (1.203–1.332) | < 0.001 |
| Coronary heart disease | OR | 1.120 (1.101–1.140) | < 0.001 | 1.422 (1.356–1.490) | < 0.001 |
| Stroke | OR | 1.080 (1.056–1.105) | < 0.001 | 1.220 (1.149–1.296) | < 0.001 |
| Heart failure | OR | 1.050 (1.015–1.086) | 0.005 | 1.296 (1.190–1.412) | < 0.001 |
| Atrial fibrillation | OR | 1.273 (1.231–1.317) | < 0.001 | 2.208 (2.044–2.385) | < 0.001 |
| Hypertension | OR | 1.152 (1.132–1.172) | < 0.001 | 1.434 (1.368–1.504) | < 0.001 |
| Kailuan community-based prospective cohort |  |  |  |  |  |
| MACCE | HR | 1.080 (1.022–1.142) | 0.006 | 1.282 (1.111–1.479) | < 0.001 |
| Stroke | HR | 1.081 (1.017–1.150) | 0.012 | 1.219 (1.038–1.432) | 0.016 |
| Heart failure | HR | 1.311 (1.124–1.527) | < 0.001 | 2.077 (1.451–2.971) | < 0.001 |
| Atrial fibrillation | HR | 1.863 (1.500–2.316) | < 0.001 | 3.923 (2.424–6.349) | < 0.001 |
| Hypertension | HR | 1.046 (1.011–1.082) | 0.010 | 1.099 (1.001–1.207) | 0.048 |

Note: CI = confidence interval; MACCE = major adverse cardiovascular and cerebrovascular events; OR = odds ratio; HR = hazard ratio. For the Tianjin Medical University Second Hospital inpatient cohort, ORs were estimated using logistic regression models. For the Kailuan community-based prospective cohort, HRs were estimated using Cox proportional hazards models. The continuous variable analysis modeled each 1-standard-deviation increase in the calibrated ECG-age gap. The high ECG-age gap group was defined as participants with a calibrated ECG-age gap z score > 1, with those having a z score ≤ 1 as the reference group. All models were adjusted for age and sex.

In the Kailuan community-based prospective cohort, the calibrated ECG-age gap also showed predictive value for future cardiovascular events. In the continuous Cox analysis, each 1-standard-deviation increase in the calibrated ECG-age gap was associated with a significantly higher risk of MACCE (HR 1.080, 95% CI 1.022–1.142). For specific outcomes, a higher calibrated ECG-age gap was significantly associated with increased risks of incident stroke, heart failure, atrial fibrillation, and hypertension. The effect sizes were most pronounced for atrial fibrillation (HR 1.863, 95% CI 1.500–2.316) and heart failure (HR 1.311, 95% CI 1.124–1.527). The categorical analysis further showed that the high ECG-age gap group had a higher risk of MACCE (HR 1.282, 95% CI 1.111–1.479), and this association was more evident for heart failure (HR 2.077, 95% CI 1.451–2.971) and atrial fibrillation (HR 3.923, 95% CI 2.424–6.349).

### Model Interpretability Analysis

To investigate the internal decision-making process of the deep learning model in physiological age prediction, we visualized the 12-lead ECG and its corresponding saliency map for a representative case (Figure 5). The subject was a female in her 50s, with an AI-predicted age in the 60s, resulting in an estimated positive age gap of approximately 11 years. In the saliency map, red regions indicate positive attribution features that drive the model to increase the predicted age, whereas blue regions represent negative attribution features that drive the model to decrease the predicted age.

**Figure 5:**
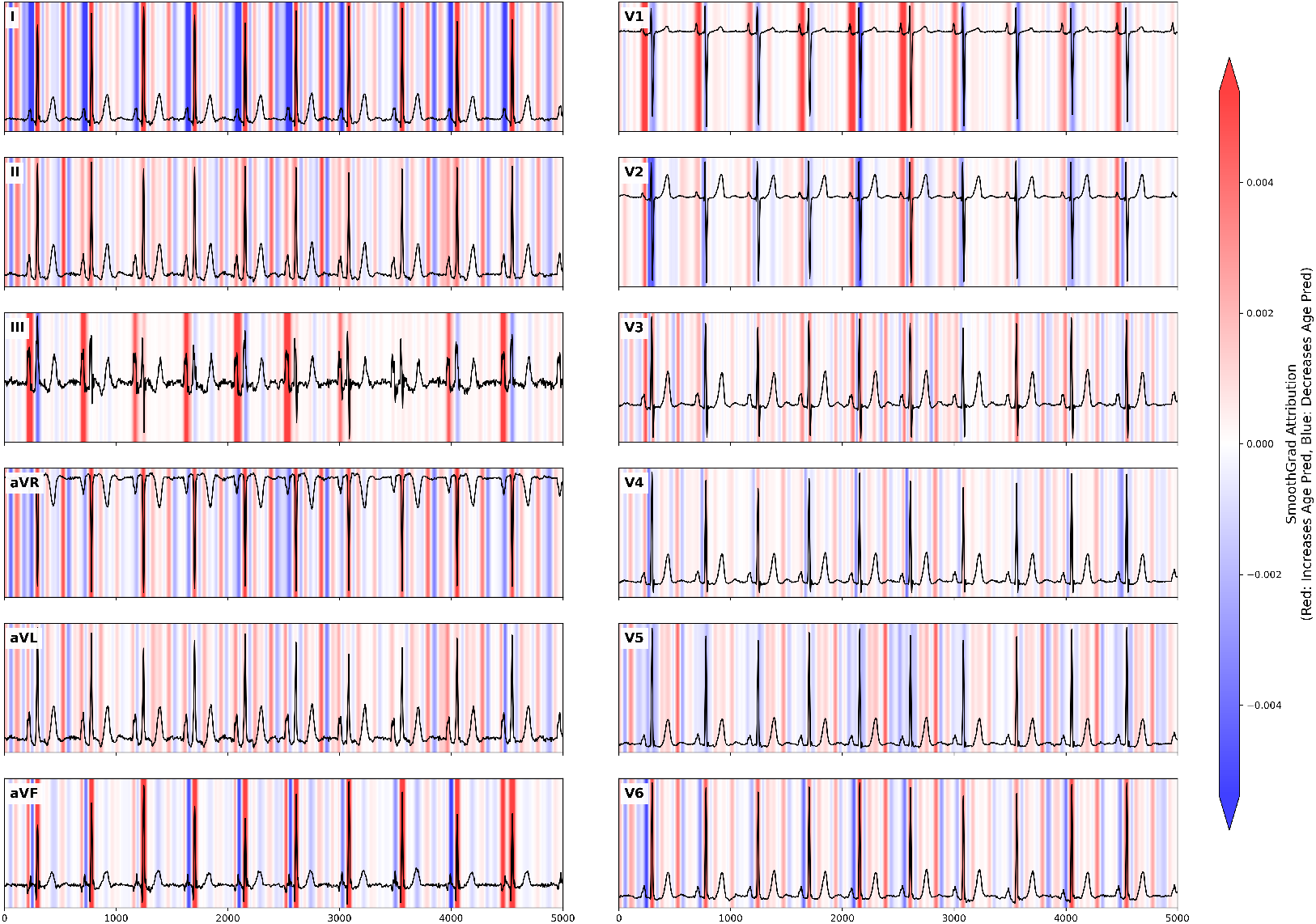
Saliency map of a representative high-risk case. Positive (red) and negative (blue) attribution features highlight the model’s primary focus on the P wave, QRS complex, and T wave, reflecting its sensitivity to subtle morphological shifts during cardiac depolarization and repolarization.

Furthermore, tightly aligned and alternating red–blue attribution bands were observed along the ascending and descending slopes of the QRS complexes as well as around their peaks and troughs. This pattern further demonstrated the model’s extreme sensitivity to subtle morphological shifts during the ventricular depolarization process.

The visualization revealed that the model did not allocate attention randomly across the background baseline. Instead, it concentrated high attribution weights on heartbeat-related regions across all leads, particularly the QRS complexes and the preceding P wave phases, with notable contributions also present around the T waves. These findings suggest that the model primarily relies on electrophysiological events during cardiac depolarization and repolarization to infer biological age. Furthermore, the presence of tightly aligned, alternating red and blue attribution bands indicated that the model did not depend on a single coarse feature but was highly sensitive to subtle morphological shifts during depolarization and repolarization.

## Discussion

In this multi-cohort study, we evaluated AI-ECG age gap as an ECG-derived digital biomarker across UK Biobank and two independent external Chinese cohorts. Using UK Biobank, we developed an AI-ECG age model by fully fine-tuning ECGFounder and demonstrated that the resulting age gap was associated with MACCE and multiple cardiovascular outcomes. Importantly, the clinical relevance of this metric was further supported in two complementary external settings. The Tianjin inpatient cohort reflected real-world hospital disease burden, whereas the Kailuan community-based prospective cohort assessed incident events in a general population setting. Across these cohorts, AI-ECG age gap showed consistent associations with cardiovascular risk and additional associations with cardiometabolic outcomes, supporting its potential as a non-invasive digital biomarker for cardiovascular aging and risk stratification.

By adopting a pre-training and fine-tuning paradigm, this study leveraged the universal electrophysiological representations learned by ECGFounder from over ten million ECGs, significantly enhancing the feature extraction efficiency for specific tasks. The introduction of the foundation model enabled the capture of subtle, aging-related electrophysiological remodeling even with a relatively limited sample size, thereby achieving robust predictions of cardiac biological age. In the experimental results, the AI-ECG predicted age and the actual calendar age showed a good linear correlation in the development set (r=0.55, MAE = 5.12 years). However, the correlation strength was moderate rather than strong, and certain prediction errors remained. We speculate that this is because the participants in the UK Biobank cohort are predominantly concentrated between 50 and 69 years old. This uneven age distribution may introduce systematic bias when the model is applied to broader populations, thereby amplifying the deviation between AI-predicted age and calendar age in the clinical cohort. On the other hand, in the clinical evaluation set, the correlation coefficient remained comparable at 0.56, and the MAE increased to 5.44 years. This may partly reflect greater deviation between AI-predicted ECG age and chronological age among individuals with cardiovascular or metabolic diseases, potentially due to accelerated cardiac aging. Despite the presence of distributional biases, the AI-ECG age gap still demonstrated a significant association with the risk of future cardiovascular events. After adjusting for age, sex, BMI, smoking history, and pre-existing comorbidities (Model 2), each 1-year increase in the age gap was still associated with a significant 13% increase in the risk of incident MACCE. Categorical analysis supported that when the AI-ECG age exceeded the calendar age by more than 6 years, the risk of MACCE was 4.51 times that of the normal population. This indicates that the aging information embedded in ECG signals captures occult cardiovascular functional degradation that is often missed in routine clinical assessments. The significant associations of the overestimation group with incident hypertension (HR = 7.10) and diabetes (HR = 6.62) suggest that the AI-ECG age gap is also related to cardiometabolic outcomes. This finding indicates that aging-related signals captured from the ECG may not solely reflect structural or functional cardiac abnormalities, but may also be influenced by metabolic dysregulation, increased vascular load, altered autonomic function, and systemic inflammation.

The two external validation cohorts further supported our findings and reflected the potential relevance of the AI-ECG age gap across different real-world settings. The inpatient cohort from Tianjin Medical University Second Hospital represented a hospital-based clinical population and was used to assess whether the AI-ECG age gap reflected existing disease burden. In this cohort, the calibrated ECG-age gap was associated with several prevalent clinical conditions, including MACCE, coronary heart disease, heart failure, atrial fibrillation, and hypertension. The Kailuan community-based prospective cohort was closer to a general population screening setting and provided longitudinal data on incident events. In this cohort, a higher calibrated ECG-age gap was associated with increased risks of future MACCE and multiple cardiovascular events. Notably, the association with incident hypertension was relatively modest, which may be related to the high incidence and multifactorial etiology of hypertension in the general population^32,33^.

The interpretability analysis provided additional support for the physiological plausibility of the model outputs. In a representative high-risk case, the saliency map showed that model attribution was concentrated on heartbeat-related components, particularly the P wave, QRS complex, and T wave, rather than being randomly distributed along the background baseline. These regions correspond to atrial depolarization, ventricular depolarization, and ventricular repolarization, respectively, and are known to be affected by age-related atrial remodeling, myocardial fibrosis or conduction system degeneration, and metabolic or autonomic changes^34–36^. These findings suggest that the AI-ECG age model may infer biological aging from physiologically meaningful ECG features related to cardiac depolarization and repolarization.

From a clinical translation perspective, AI-ECG age prediction relies only on routine ECG examination and has advantages such as low cost, non-invasiveness, high reproducibility, and broad accessibility. It therefore has potential for real-world application. This tool could be integrated into existing routine health examination workflows or primary care screening systems to help identify high-risk individuals at the subclinical stage, thereby providing reference information for early intervention and long-term health management.

However, this study still has certain limitations. First, addressing the prediction bias caused by the uneven age distribution observed in this study, future algorithms still need optimization. For instance, incorporating a distribution-aware loss function to mitigate regression shift caused by sample imbalance^37^, or adopting a post-hoc statistical calibration strategy to align the distribution of predicted values^30,38^, could enhance the prediction consistency and accuracy across the entire age spectrum. Furthermore, future studies should integrate longitudinal follow-up data with multi-omics profiling to characterize the biological mechanisms underlying cardiac electrical aging acceleration and to evaluate the feasibility of AI-ECG age as a metric for assessing the effectiveness of lifestyle interventions.

Despite these limitations, the present study provides supportive evidence that AI-ECG age, as a highly scalable and non-invasive digital biomarker, possesses substantial potential in the cardiovascular and cardiometabolic risk stratification of broad populations. With the continuous advancement of artificial intelligence technologies and the increasing adoption of electrocardiographic monitoring devices, AI-ECG age is expected to become a useful tool in next-generation digital preventive cardiology, providing crucial support for the formulation of public health strategies and clinical decision-making.

## Data Availability

The UK Biobank data used in this study are available through application to the UK Biobank under approved access procedures (Application No. 90018). The external validation data from Tianjin Medical University Second Hospital and the Kailuan community-based prospective cohort are not publicly available because of participant privacy considerations and institutional ethical restrictions.

https://www.ukbiobank.ac.uk/use-our-data/apply-for-access/

## Acknowledgments

This work is supported by Beijing Municipal Science and Technology Commission (Z25110000072 5008), the National Natural Science Foundation of China (62102008), CCF-Tencent Rhino-Bird Open Research Fund (CCF-Tencent RAGR20250108), CCF-Zhipu Large Model Innovation Fund (CCF-Zhipu202414), PKU-OPPO Fund (BO202301, BO202503), Research Project of Peking University in the State Key Laboratory of Vascular Homeostasis and Remodeling (2025-SKLVHR-YCTS-02), Prevention and Control of Emerging and Major Infectious Diseases–National Science and Technology Major Project (2025ZD01906000, 2025ZD01906004).

## Data and Code Availability

The UK Biobank data used in this study are available through application to the UK Biobank under approved access procedures (Application No. 90018). The external validation data from Tianjin Medical University Second Hospital and the Kailuan community-based prospective cohort are not publicly available because of participant privacy considerations and institutional ethical restrictions. The source code and model implementation for the AI-ECG age prediction framework are publicly available at: https://github.com/PKUDigitalHealth/ECG-age.

## Appendix Supplementary Figures

**Figure S1:**
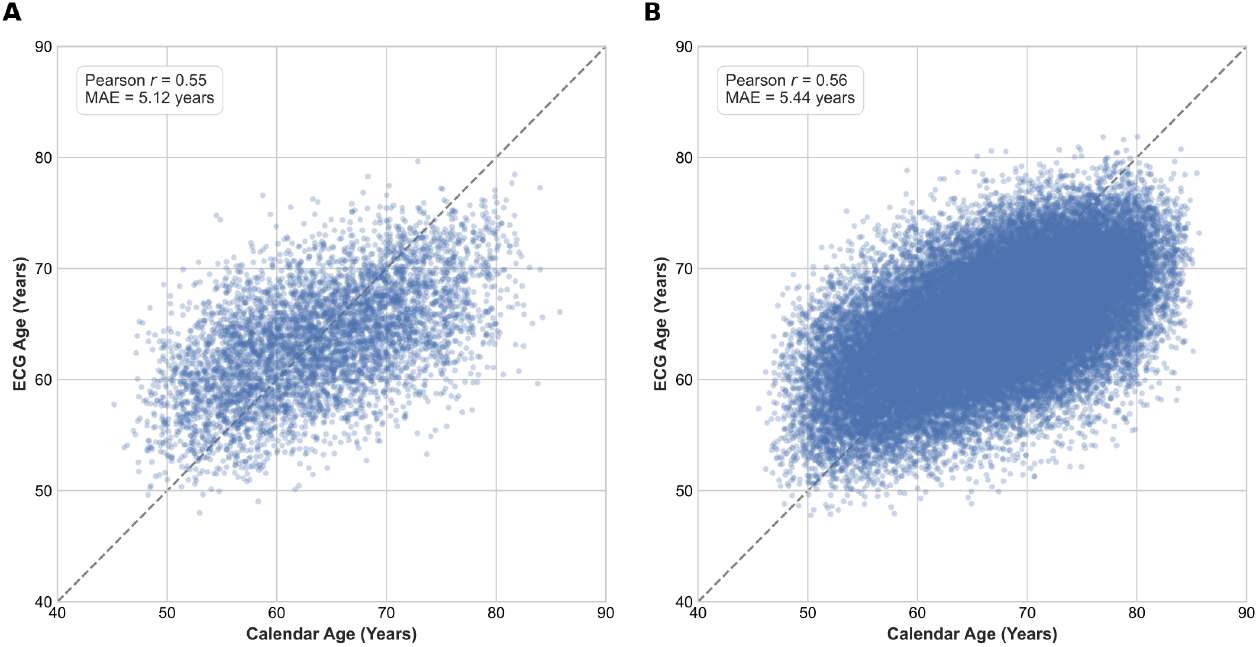
Scatter plots demonstrating the correlation between calendar age and AI-ECG predicted age in the internal cohorts. The plots show the prediction consistency in the development cohort (test set) (r = 0.55, MAE = 5.12 years) and the clinical evaluation cohort (r = 0.56, MAE = 5.44 years).

## References

1. Naghavi, M., Ong, K.L., Aali et al. (2024). Global burden of 288 causes of death and life expectancy decomposition in 204 countries and territories and 811 subnational locations, 1990–2021: a systematic analysis for the global burden of disease study 2021. The Lancet 403, 2100–2132. URL: http://dx.doi.org/10.1016/S0140-6736(24)00367-2. doi: 10.1016/s0140-6736(24)00367-2.

2. Stark, B.A., DeCleene, N.K., Desai et al. (2025). Global, regional, and national burden of cardiovascular diseases and risk factors in 204 countries and territories, 1990-2023. JACC 86, 2167–2243. URL: http://dx.doi.org/10.1016/j.jacc.2025.08.015. doi: 10.1016/j.jacc.2025.08.015.

3. World Health Organization (2025). Cardiovascular diseases (cvds) fact sheet.. URL: https://www.who.int/news-room/fact-sheets/detail/cardiovascular-diseases-(cvds).

4. Brauer, M., Roth, G.A., Aravkin et al. (2024). Global burden and strength of evidence for 88 risk factors in 204 countries and 811 subnational locations, 1990–2021: a systematic analysis for the global burden of disease study 2021. The Lancet 403, 2162–2203. URL: http://dx.doi.org/10.1016/S0140-6736(24)00933-4. doi: 10.1016/s0140-6736(24)00933-4.

5. van Daalen, K.R., Zhang, D., Kaptoge et al. (2024). Risk estimation for the primary prevention of cardiovascular disease: considerations for appropriate risk prediction model selection. The Lancet Global Health 12, e1343–e1358. URL: http://dx.doi.org/10.1016/S2214-109X(24)00210-9. doi: 10.1016/s2214-109x(24)00210-9.

6. Krittanawong, C., Rogers, A.J., Johnson et al. (2020). Integration of novel monitoring devices with machine learning technology for scalable cardiovascular management. Nature Reviews Cardiology 18, 75–91. URL: http://dx.doi.org/10.1038/s41569-020-00445-9. doi: 10.1038/s41569-020-00445-9.

7. Iyngkaran, P., Appuhamilage, P.Y., Patabandige et al. (2024). Barriers to cardiac rehabilitation among patients diagnosed with cardiovascular diseases—a scoping review. International Journal of Environmental Research and Public Health 21, 339. URL: http://dx.doi.org/10.3390/ijerph21030339. doi: 10.3390/ijerph21030339.

8. Siontis, K.C., Noseworthy, P.A., Attia et al. (2021). Artificial intelligence-enhanced electrocardiography in cardiovascular disease management. Nature Reviews Cardiology 18, 465–478. URL: http://dx.doi.org/10.1038/s41569-020-00503-2. doi: 10.1038/s41569-020-00503-2.

9. Yang, K., Hong, M., Zhang, J. et al. (2025). Ecg-lm: Understanding electrocardiogram with a large language model. Health Data Science 5. URL: http://dx.doi.org/10.34133/hds.0221. doi: 10.34133/hds.0221.

10. Huang, S., Zhang, D., Fan, S. et al. (2025). Opportunistic screening of wolff-parkinson-white syndrome using single-lead ai-ecg mobile system a real-world study of over 3.5 million ecg recordings in china. arXiv preprint arXiv:2510.24750.

11. Zhao, Q., Geng, S., Wang, B. et al. (2024). Deep learning in heart sound analysis: From techniques to clinical applications. Health Data Science 4. URL: http://dx.doi.org/10.34133/hds.0182. doi: 10.34133/hds.0182.

12. Huang, S., Xing, W., Geng, S. et al. (2025). Combining ecg foundation model and xg-boost to predict in-hospital malignant ventricular arrhythmias in ami patients. arXiv preprint arXiv:2510.17172.

13. Zhang, S., Mu, W., Dong, D. et al. (2023). The applications of artificial intelligence in digestive system neoplasms: A review. Health Data Science 3. URL: http://dx.doi.org/10.34133/hds.0005. doi: 10.34133/hds.0005.

14. Li, J., Aguirre, A.D., Junior, V.M. et al. (2025). An electrocardiogram foundation model built on over 10 million recordings. NEJM AI 2. URL: http://dx.doi.org/10.1056/aioa2401033. doi: 10.1056/aioa2401033.

15. Tao, Y., Zhang, D., Pang, N. et al. (2024). Multi-modal artificial intelligence algorithm for the prediction of left atrial low-voltage areas in atrial fibrillation patient based on sinus rhythm electrocardiogram and clinical characteristics: a retrospective, multicentre study. European Heart Journal - Digital Health 6, 200–208. URL: http://dx.doi.org/10.1093/ehjdh/ztae095. doi: 10.1093/ehjdh/ztae095.

16. Fiorina, L., Carbonati, T., Narayanan, K. et al. (2025). Near-term prediction of sustained ventricular arrhythmias applying artificial intelligence to single-lead ambulatory electrocardiogram. European Heart Journal 46, 1998–2008. URL: http://dx.doi.org/10.1093/eurheartj/ehaf073. doi: 10.1093/eurheartj/ehaf073.

17. Ziegler, K.A., Engelhardt, S., Carnevale, D. et al. (2025). Neural mechanisms in cardiovascular health and disease. Circulation Research 136, 1233–1261. URL: http://dx.doi.org/10.1161/CIRCRESAHA.125.325580. doi: 10.1161/circresaha.125.325580.

18. Bao, X., Zhang, Z., Shen, Y. et al. (2025). Cardiovascular information and health engineering medicine. Research 8. URL: http://dx.doi.org/10.34133/research.0956. doi: 10.34133/research.0956.

19. Ritchie, R.H., and Abel, E.D. (2020). Basic mechanisms of diabetic heart disease. Circulation Research 126, 1501–1525. URL: http://dx.doi.org/10.1161/CIRCRESAHA.120.315913. doi: 10.1161/circresaha.120.315913.

20. Nie, G., Zhao, Q., Tang, G. et al. (2025). Artificial intelligence-derived photoplethysmography age as a digital biomarker for cardiovascular health. Communications Medicine 5. URL: http://dx.doi.org/10.1038/s43856-025-01188-9. doi: 10.1038/s43856-025-01188-9.

21. Zhang, D., Li, J., Geng, S. et al. (2026). ECGomics an open platform for AI-ECG digital biomarker discovery. arXiv preprint arXiv:2601.15326.

22. Chen, H., Cao, Z., Zhang, J. et al. (2025). Accelerometer-measured physical activity and neuroimaging-driven brain age. Health Data Science 5. URL: http://dx.doi.org/10.34133/hds.0257. doi: 10.34133/hds.0257.

23. Lima, E.M., Ribeiro, A.H., Paixão, G.M.M. et al. (2021). Deep neural network-estimated electrocardiographic age as a mortality predictor. Nature Communications 12. URL: http://dx.doi.org/10.1038/s41467-021-25351-7. doi: 10.1038/s41467-021-25351-7.

24. Cho, S., Eom, S., Kim, D. et al. (2024). Artificial intelligence–derived electrocardiographic aging and risk of atrial fibrillation: a multi-national study. European Heart Journal 46, 839–852. URL: http://dx.doi.org/10.1093/eurheartj/ehae790. doi: 10.1093/eurheartj/ehae790.

25. Hempel, P., Ribeiro, A.H., Vollmer, M. et al. (2025). Explainable ai associates ecg aging effects with increased cardiovascular risk in a longitudinal population study. npj Digital Medicine 8. URL: http://dx.doi.org/10.1038/s41746-024-01428-7. doi: 10.1038/s41746-024-01428-7.

26. Attia, Z.I., Friedman, P.A., Noseworthy, P.A. et al. (2019). Age and sex estimation using artificial intelligence from standard 12-lead ecgs. Circulation: Arrhythmia and Electrophysiology 12. URL: http://dx.doi.org/10.1161/CIRCEP.119.007284. doi: 10.1161/circep.119.007284.

27. Sudlow, C., Gallacher, J., Allen, N. et al. (2015). Uk biobank: An open access resource for identifying the causes of a wide range of complex diseases of middle and old age. PLOS Medicine 12, e1001779. URL: http://dx.doi.org/10.1371/journal.pmed.1001779. doi: 10.1371/journal.pmed.1001779.

28. He, K., Zhang, X., Ren, S. et al. (2016). Deep residual learning for image recognition. In Proceedings of the IEEE conference on computer vision and pattern recognition. pp. 770–778.

29. Hong, S., Xu, Y., Khare, A. et al. (2020). Holmes: Health online model ensemble serving for deep learning models in intensive care units. In Proceedings of the 26th ACM SIGKDD International Conference on Knowledge Discovery & Data Mining. KDD’20. ACM pp. 1614–1624. URL: http://dx.doi.org/10.1145/3394486.3403212. doi: 10.1145/3394486.3403212.

30. Le Goallec, A., Diai, S., Collin, S., Prost, J.B., Vincent, T., and Patel, C.J. (2022). Using deep learning to predict abdominal age from liver and pancreas magnetic resonance images. Nature Communications 13. URL: http://dx.doi.org/10.1038/s41467-022-29525-9. doi: 10.1038/s41467-022-29525-9.

31. Barthels, M., Verhofstadt, E., Delgado, I.B., Gruwez, H., Pison, L., Pierlet, N., and Vandervoort, P. (2025). Artificial intelligence-predicted ecg age gap as a biomarker: bias-adjusted correlation with mortality and cardiovascular risk factors. European Heart Journal - Digital Health 7. URL: http://dx.doi.org/10.1093/ehjdh/ztaf137. doi: 10.1093/ehjdh/ztaf137.

32. Mills, K.T., Stefanescu, A., and He, J. (2020). The global epidemiology of hypertension. Nature Reviews Nephrology 16, 223–237. URL: http://dx.doi.org/10.1038/s41581-019-0244-2. doi: 10.1038/s41581-019-0244-2.

33. Zhao, H.Y., Liu, X.X., Wang, A.X., Wu, Y.T., Zheng, X.M., Zhao, X.H., Cui, K., Ruan, C.Y., Lu, C.Z., Jonas, J.B., and Wu, S.L. (2016). Ideal cardiovascular health and incident hypertension: The longitudinal community-based kailuan study. Medicine 95, e5415. URL: http://dx.doi.org/10.1097/MD.0000000000005415. doi: 10.1097/md.0000000000005415.

34. Magnani, J.W., Williamson, M.A., Ellinor, P.T. et al. (2009). P wave indices: Current status and future directions in epidemiology, clinical, and research applications. Circulation: Arrhythmia and Electrophysiology 2, 72–79. URL: http://dx.doi.org/10.1161/CIRCEP.108.806828. doi: 10.1161/circep.108.806828.

35. Lakatta, E.G., and Levy, D. (2003). Arterial and cardiac aging: Major shareholders in cardiovascular disease enterprises: Part i: Aging arteries: A “set up” for vascular disease. Circulation 107, 139–146. URL: http://dx.doi.org/10.1161/01.cir.0000048892.83521.58. doi: 10.1161/01.cir.0000048892.83521.58.

36. Chen, P.S., Chen, L.S., Fishbein, M.C. et al. (2014). Role of the autonomic nervous system in atrial fibrillation: Pathophysiology and therapy. Circulation Research 114, 1500–1515. URL: http://dx.doi.org/10.1161/CIRCRESAHA.114.303772. doi: 10.1161/circresaha.114.303772.

37. Nie, G., Tang, G., and Hong, S. (2025). Dist loss: Enhancing regression in few-shot region through distribution distance constraint. In The Thirteenth International Conference on Learning Representations. URL: https://openreview.net/forum?id=YeSxbRrDRl.

38. Le, T.T., Kuplicki, R.T., McKinney, B.A. et al. (2018). A nonlinear simulation framework supports adjusting for age when analyzing brainage. Frontiers in Aging Neuroscience 10. URL: http://dx.doi.org/10.3389/fnagi.2018.00317. doi: 10.3389/fnagi.2018.00317.

